# Effectiveness of slow-release oral morphine versus other OAT regimens in key sub-populations: protocol for population-based target trial emulation

**DOI:** 10.64898/2025.11.30.25341316

**Authors:** Momenul Haque Mondol, Jeong Eun Min, Michelle Zanette, Maria Eugenia Socías, Paul Gustafson, Paxton Bach, Robert William Platt, Shaun Seaman, Sander Greenland, Bohdan Nosyk

**Affiliations:** School of Population and Public Health, University of British Columbia, Vancouver, Canada; Centre for Advancing Health Outcomes, Vancouver, British Columbia, Canada; Department of Statistics and Data Science, University of Barishal, Barishal, Bangladesh; Department of Medicine, University of British Columbia, Vancouver, British Columbia, Canada; British Columbia Centre on Substance Use, Vancouver, British Columbia, Canada; Department of Statistics, University of British Columbia, Vancouver, British Columbia, Canada; Department of Epidemiology, Biostatistics and Occupational Health, McGill University, Montreal, Quebec, Canada; MRC Biostatistics Unit, Cambridge, UK; Department of Epidemiology and Department of Statistics, UCLA, Los Angeles, California, USA; Faculty of Health Sciences, Simon Fraser University; Burnaby, British Columbia, Canada

## Abstract

**Introduction:** Slow-release oral morphine (SROM) was introduced as an alternative form of opioid agonist treatment (OAT) in British Columbia (BC), Canada in 2017. While clinical guidelines in BC recommend SROM based on expert consensus and experience, there is limited real-world evidence on populations most likely to benefit from SROM compared to other forms of OAT, particularly in the context of widespread fentanyl exposure. We will estimate the comparative effectiveness of SROM versus methadone and buprenorphine/naloxone on OAT discontinuation and all-cause mortality among key sub-populations in BC.

**Methods:** We will conduct a population-level retrospective cohort study using linked data from nine provincial health administrative databases. The study population includes adults (≥18 years) in BC who initiated SROM, methadone, or buprenorphine/naloxone between June 1, 2017, and December 31, 2022. Key sub-populations will include incident users with no prior OAT history and prevalent new users with prior OAT experience stratified by OAT type, stability, and medication switching history. The primary outcomes are time to OAT discontinuation and all-cause mortality, while overdose-related acute care visits will be examined as a secondary outcome. To estimate both the initiator effect and the effect of treatment at guideline-recommended doses, we will apply marginal structural models using inverse probability treatment weighting and ‘clone-censor-weights’ approach to address confounding by indication and time-varying confounding. Sensitivity analyses will evaluate the robustness of our findings, including cohort and timeline restrictions, alternative outcome definitions, and alternative estimation strategies including high-dimensional propensity score and instrumental variable approaches.

**Discussion:** This study will generate real-world evidence on the comparative effectiveness of SROM versus methadone and buprenorphine/naloxone across clinically distinct OAT sub-populations. The findings will support evidence-informed updates to OAT guidelines and clinical decision-making in BC and other jurisdictions facing escalating opioid-related harms.

## Introduction

### Slow-release oral morphine as an alternative OAT

Methadone and buprenorphine/naloxone are common pharmacological treatments for opioid use disorder (OUD) aimed at reducing unregulated opioid use, overdose risk, and associated harms.[1–10] However, not all individuals respond well to these treatments, necessitating alternative options such as slow-release oral morphine (SROM), extended-release naltrexone, extended-release buprenorphine, and injectable opioid agonist therapies.[6,8,11] Methadone, a full mu-opioid receptor agonist, has long been the gold standard for OAT due to its high retention rates but requires careful dose titration due to its long half-life, overdose risk, and potential for QT (a measure of the heart’s electrical cycle on electrocardiogram) prolongation.[12] Buprenorphine/naloxone, a partial agonist, has a better safety profile but exhibits a ceiling effect on opioid receptor activation, which may limit its efficacy for individuals with high opioid tolerance.[6]

In contrast, SROM is a full agonist with a long-acting formulation which provides stable plasma levels with once-daily dosing without a ceiling on its opioid effects, making it a potential alternative to methadone and buprenorphine/naloxone. Additionally, unlike methadone, SROM does not prolong the QT interval and may have a lower risk of sedation.[13–15] Originally developed for pain management, SROM has gained recognition as an opioid agonist treatment (OAT) in several countries, including Austria, Germany, and Canada, though its regulatory approval status varies. SROM, marketed as Kadian^®^, a 24-hour extended-release morphine capsule, was introduced in 2017 under British Columbia, Canada’s (BC) provincial drug plan (PharmaCare) as an off-label option for individuals who have not found success with methadone or buprenorphine/naloxone, though it is unclear who are most likely to benefit.[5,6]

### Adapting clinical guidelines to a new drug landscape

Clinical guidelines in BC had previously regarded SROM as a third-line treatment behind buprenorphine/naloxone and methadone but updated their guidance in 2023, in response to rising overdose death rates across the province, to remove the tiers and advise collaborative treatment selection based on client preferences, clinical judgement and contextual circumstances.[6] Guidance for SROM is largely based on clinical consensus and expert opinion due to overall limited evidence to support decision-making.[6] This client-centered shift acknowledges that prior experiences with OAT often shape future treatment decisions. SROM may be an alternative option for individuals who have experienced negative outcomes with other OAT medications, such as incomplete elimination of withdrawal with buprenorphine/naloxone, or intolerable side effects with methadone, including sedation, emotional blunting, or cardiac risks such as QT prolongation.[11] Individuals with contraindications to methadone, including ventricular arrhythmias or concurrent use of QT-prolonging medications, may particularly benefit from SROM as the medication does not affect the QT interval.[11] Moreover, frequent or early switching between OAT options is often a signal of suboptimal treatment fit, whether due to poor tolerability, inadequate therapeutic response, or individual treatment goals not being met.[6,11]

While the 2023 BC guideline outlined expert-informed protocols for switching from methadone or buprenorphine to SROM, these recommendations are not supported by empirical evidence, and the comparative effectiveness of this form of OAT remains uncertain. Moreover, the unregulated drug supply in BC has been increasingly contaminated with fentanyl, a synthetic opioid 40–100 times more potent than heroin. By 2017, fentanyl was detected in over 80% of unregulated drug deaths in the province.[16] People who use fentanyl may have higher tolerance and require higher doses to maintain OAT compared to those using prescription opioids, highlighting the urgent need for tailored treatment approaches.[17] This shift challenges existing OAT guidelines as they are based on evidence established before fentanyl was a dominant drug in the unregulated supply. Given fentanyl’s increasing prevalence and role in rising overdose risks, it is critical that we have an understanding of the comparative effectiveness and best practices for every OAT option in fentanyl-tolerant individuals.

### Evidence on SROM’s effectiveness and safety

Some studies suggest that SROM offers comparable retention rates to methadone, with potential advantages in reducing heroin cravings, improving mental health outcomes, and increasing individual’s treatment satisfaction.[18–21] However, others raised concerns regarding shortage of evidence, misuse, and lack of standardized guidelines.[6,22] A 2010 systematic review and a 2013 Cochrane review found that SROM was comparable to methadone in retention rates and withdrawal symptom relief, with potential benefits in reducing depression and cravings.[18,19] However, the former reviewed 13 studies involving a total of 750 individuals, of which only one was a randomized controlled trial (RCT) with 64 participants followed for 7 weeks, while eight studies lacked a comparison group.[18] The Cochrane review included only three RCTs with a total of 195 participants, of which two were short-duration crossover trials lasting 6 and 7 weeks, and none enrolled individuals with documented poor response to other maintenance treatments.[19] A 2019 meta-analysis of four RCTs (including the same three studies from the 2013 Cochrane review) reinforced these findings, indicating SROM’s equivalence to methadone in retention and heroin use reduction.[23] However, all participants were recruited from European settings before 2010 and were stabilized on OAT, limiting generalizability to more clinically complex populations.

Another meta-analysis was conducted in 2022 to investigate the effectiveness of different OAT medications, and SROM was only featured in 4 of 79 RCTs.[24] As a result, the authors could not draw conclusions regarding SROM’s comparative effectiveness on treatment retention due to limited high-quality trials. Notably, only 6 distinct RCTs that compare SROM to another OAT medication were included among these systematic reviews and meta-analyses, with the most recent trial conducted over 10 years ago. None of these RCTs were conducted among people who use illicit fentanyl or had indicated a poor response to another OAT medication prior to the trial with SROM,[23,24] and only one trial compared SROM with buprenorphine.[25]

More recently, the pRESTO trial in Vancouver (2021) aimed to evaluate SROM’s effectiveness compared to methadone in fentanyl-exposed populations but was terminated early due to COVID-19, leaving significant gaps in understanding its long-term retention and safety in high-risk patients.[26,27] Observational studies conducted in Germany in 2016 and 2017 showed successful transitions from methadone and buprenorphine to SROM,[28] 60.6% retention at 1 year, reduction in heroin and intravenous drug use, and important improvements in mental and physical health among patients with extensive, but often unsatisfactory prior OAT experience.[29] Additionally, a 2021 hospital-based study in Canada found the ability to offer SROM, in addition to methadone and buprenorphine, resulted in increased rates of OAT initiation among hospitalized patients with OUD, high post-discharge retention, and that patients with chronic pain were more likely to initiate with SROM.[30] However, these studies were limited by small sample sizes[30] and did not compare long-term SROM retention to methadone or buprenorphine.[29,30]

Concerns regarding SROM’s safety have raised questions about its role in OAT. A 2016 epidemiological study in France reported high rates of SROM use by injection, and illegal acquisition, indicating significant risks of misuse and diversion.[31] Also in France, a 2020 population-based cohort study (using data from 2012-2014) found that SROM users had 3.8-fold and 2-fold higher overdose incidence and 9.1-fold and 3.9-fold higher mortality incidence compared to buprenorphine and methadone, respectively, along with increased bacterial infections. However, a relatively small cohort of SROM users had more comorbidities such as mental health and alcohol use disorders but were prescribed benzodiazepines less than methadone or buprenorphine users, indicating less comprehensive care than those receiving these conventional treatments and potential selection bias.[32] Nonetheless, evaluating the real-world effectiveness of SROM as second-line treatment, compared to methadone and buprenorphine/naloxone, among key sub-populations and determining who will benefit most from this form of OAT remains a pressing public health concern amid the ongoing overdose crisis.[22,33]

### Impact of COVID-19 on OAT policies and outcomes

The COVID-19 pandemic necessitated substantial changes in OAT delivery to ensure continued care while minimizing in-person interactions. Across multiple jurisdictions, including Canada,[34,35] the U.S.,[36,37] Australia,[38] New Zealand,[39] and the United Kingdom,[40] regulatory changes were implemented to expand take-home dosing, integrate telehealth consultations, and ease urine drug testing requirements. These policy shifts broadened OAT accessibility, particularly for individuals facing barriers to in-person daily treatment.[35] A survey-based study from Ontario involving 402 patients and 100 prescribers found that expanded unsupervised dosing during COVID-19 was not associated with increased rates of overdose, emergency department visits, or hospitalizations, although diversion behaviors such as dose sharing and trading were more frequently reported.[35] While these adaptations largely applied to methadone and buprenorphine/naloxone, the impact on SROM accessibility and treatment outcomes remains unclear.[34] Interviews with 25 opioid treatment program directors in the U.S. revealed mostly positive responses to COVID-related regulatory changes, though some raised concerns about reduced clinical oversight, medication diversion, and patient isolation.[41] Additionally, while 37 U.S. states initially adopted expanded methadone take-home flexibilities, four states later rescinded these changes, reverting to pre-pandemic regulations.[37] These findings highlight the variability in policy implementation across jurisdictions and raise questions about the long-term sustainability of these regulatory adaptations.

BC operates within a single-payer healthcare system where access to OAT is not restricted by site type or insurance status. Unlike the U.S., where OAT coverage often depends on Medicaid expansion and site-specific regulations (e.g., methadone often limited to certified clinics), BC allows OAT delivery in diverse clinical settings and provides a wider range of medications, including SROM. This unique regulatory landscape provides a rare opportunity to generate real-world evidence on SROM’s effectiveness relative to methadone and buprenorphine/naloxone using the population-based linked administrative dataset in BC.

### Objective

Our objective is to conduct a population-level retrospective observational study using nine linked provincial administrative databases to compare the effectiveness of SROM versus methadone and buprenorphine/naloxone in BC from 2017 to 2022 among key sub-populations. We will evaluate both the initiator effect (treatment outcomes based on initial OAT dispensation) and the per-protocol effect (treatment outcomes when adhering to the initial OAT regimen, according to BC clinical guidelines) on treatment discontinuation and all-cause mortality in real-world settings.

## Materials and Methods

### Study design

This study is designed as a population-based retrospective cohort study, aiming to emulate a suite of target trials (i.e., hypothetical randomized clinical trials, RCTs) for individuals initiating methadone, buprenorphine/naloxone, or SROM between June 1, 2017, and December 31, 2022, in BC, Canada. This timeline ensures the availability of SROM alongside methadone and buprenorphine/naloxone in BC. Although RCTs are the gold standard for determining comparative effectiveness, they are often time-intensive and may lack generalizability to broader population.[42] By leveraging large-scale health administrative datasets and advanced statistical techniques, observational studies can address many common biases and yield treatment effect estimates based on real-world evidence.[43–46] We will explore the effect variation across key sub-populations which will help identify most beneficiary group from SROM compared to methadone, buprenorphine/naloxone. Key elements of the target trials and their emulations are summarized in **Table 1**.

**Table 1:**
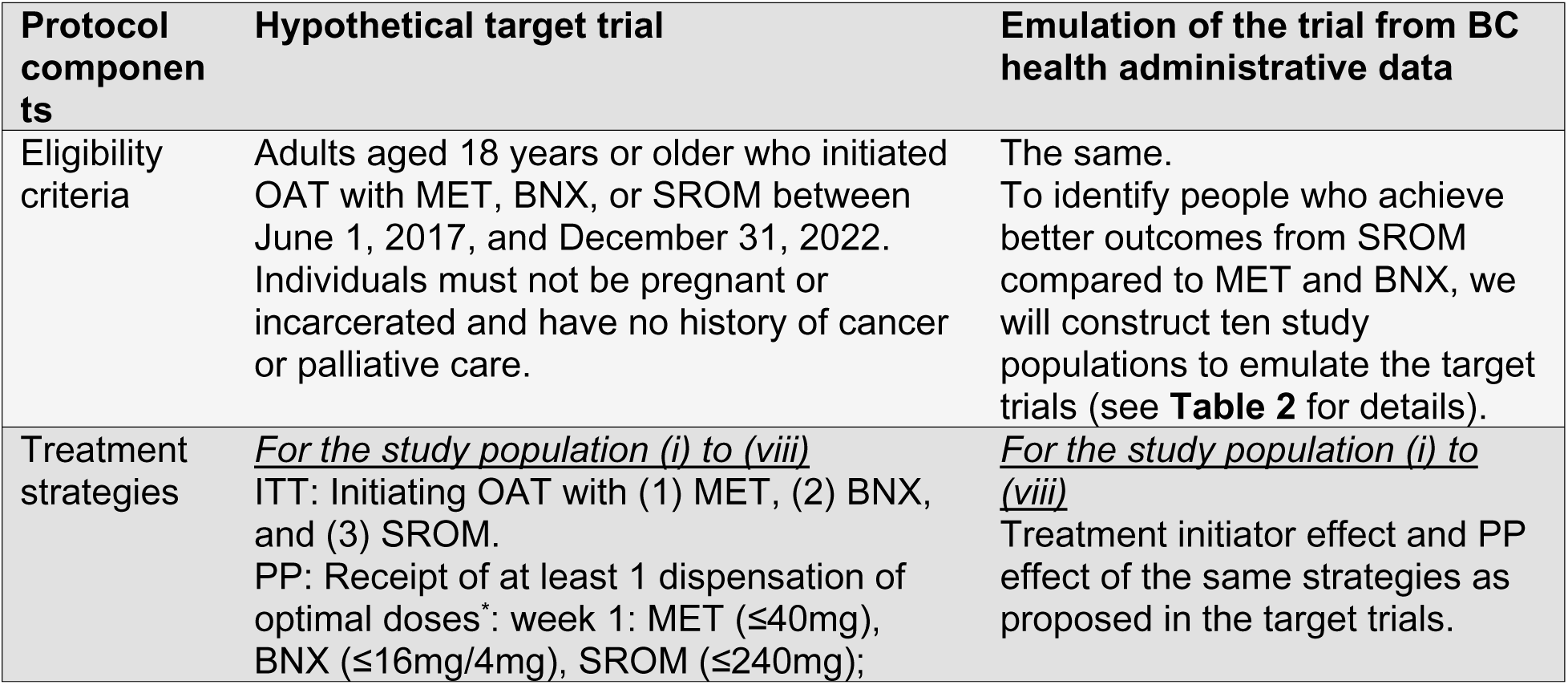
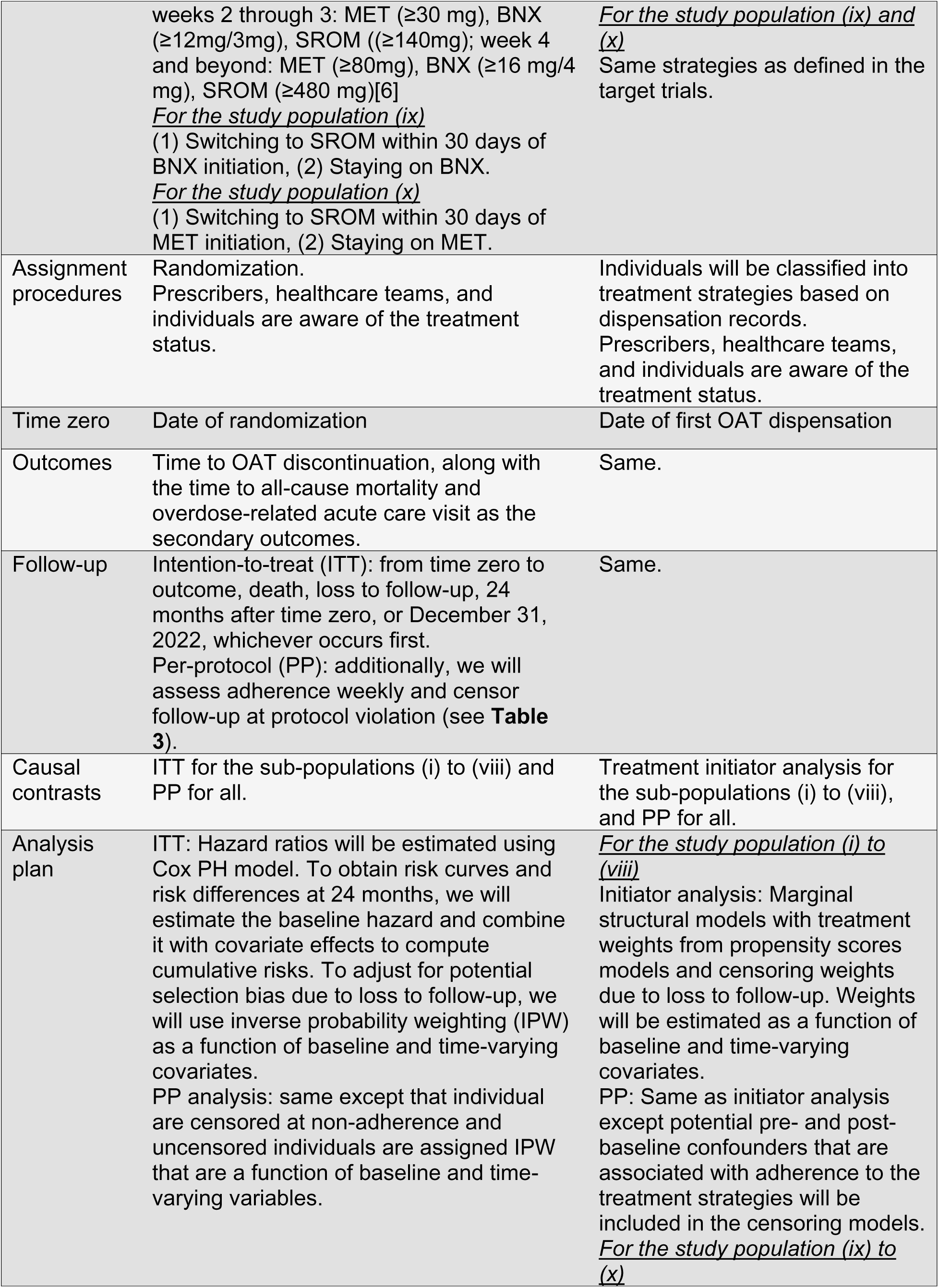
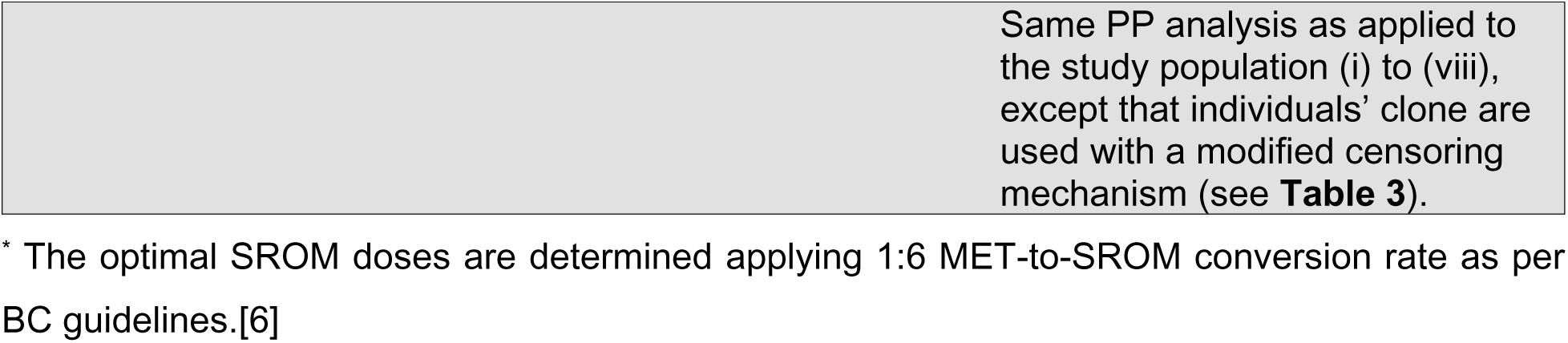
Key components of the emulated target trials on comparing the initiation of SROM versus methadone and buprenorphine/naloxone to treat people with opioid use disorder using the observational data.

| Protocol components | Hypothetical target trial | Emulation of the trial from BC health administrative data |
| --- | --- | --- |
| Eligibility criteria | Adults aged 18 years or older who initiated OAT with MET, BNX, or SROM between June 1, 2017, and December 31, 2022. Individuals must not be pregnant or incarcerated and have no history of cancer or palliative care. | The same.<br>To identify people who achieve better outcomes from SROM compared to MET and BNX, we will construct ten study populations to emulate the target trials (see **Table 2** for details). |
| Treatment strategies | <u>For the study population (i) to (viii)</u><br>ITT: Initiating OAT with (1) MET, (2) BNX, and (3) SROM.<br>PP: Receipt of at least 1 dispensation of optimal doses*: week 1: MET ($\leq 40\text{mg}$), BNX ($\leq 16\text{mg}/4\text{mg}$), SROM ($\leq 240\text{mg}$); | <u>For the study population (i) to (viii)</u><br>Treatment initiator effect and PP effect of the same strategies as proposed in the target trials. |
|  | weeks 2 through 3: MET ($\geq 30$ mg), BNX ($\geq 12$mg/3mg), SROM ($\geq 140$mg); week 4 and beyond: MET ($\geq 80$mg), BNX ($\geq 16$ mg/4 mg), SROM ($\geq 480$ mg)[6]<br><u>For the study population (ix)</u><br>(1) Switching to SROM within 30 days of BNX initiation, (2) Staying on BNX.<br><u>For the study population (x)</u><br>(1) Switching to SROM within 30 days of MET initiation, (2) Staying on MET. | <u>For the study population (ix) and (x)</u><br>Same strategies as defined in the target trials. |
| Assignment procedures | Randomization.<br>Prescribers, healthcare teams, and individuals are aware of the treatment status. | Individuals will be classified into treatment strategies based on dispensation records.<br>Prescribers, healthcare teams, and individuals are aware of the treatment status. |
| Time zero | Date of randomization | Date of first OAT dispensation |
| Outcomes | Time to OAT discontinuation, along with the time to all-cause mortality and overdose-related acute care visit as the secondary outcomes. | Same. |
| Follow-up | Intention-to-treat (ITT): from time zero to outcome, death, loss to follow-up, 24 months after time zero, or December 31, 2022, whichever occurs first.<br>Per-protocol (PP): additionally, we will assess adherence weekly and censor follow-up at protocol violation (see **Table 3**). | Same. |
| Causal contrasts | ITT for the sub-populations (i) to (viii) and PP for all. | Treatment initiator analysis for the sub-populations (i) to (viii), and PP for all. |
| Analysis plan | ITT: Hazard ratios will be estimated using Cox PH model. To obtain risk curves and risk differences at 24 months, we will estimate the baseline hazard and combine it with covariate effects to compute cumulative risks. To adjust for potential selection bias due to loss to follow-up, we will use inverse probability weighting (IPW) as a function of baseline and time-varying covariates.<br>PP analysis: same except that individual are censored at non-adherence and uncensored individuals are assigned IPW that are a function of baseline and time-varying variables. | <u>For the study population (i) to (viii)</u><br>Initiator analysis: Marginal structural models with treatment weights from propensity scores models and censoring weights due to loss to follow-up. Weights will be estimated as a function of baseline and time-varying covariates.<br>PP: Same as initiator analysis except potential pre- and post-baseline confounders that are associated with adherence to the treatment strategies will be included in the censoring models.<br><u>For the study population (ix) to (x)</u> |

|  |  |
| --- | --- |
|  | Same PP analysis as applied to the study population (i) to (viii), except that individuals' clone are used with a modified censoring mechanism (see **Table 3**). |

\* The optimal SROM doses are determined applying 1:6 MET-to-SROM conversion rate as per BC guidelines.[6]

This study will utilize population-level data derived from the linkage of nine provincial health administrative databases in BC, where residents are required to participate in a single-payer health insurance system. Using the BC PharmaNet database,[47] which captures daily medication dispensations, we will identify dispensation dates and doses for methadone, buprenorphine/naloxone, and SROM (refer to **S1 Table** for a list of drug/product identification numbers) to define treatment episode. Additional databases include the Discharge Abstract Database[48] (DAD, documenting hospitalizations), Medical Services Plan[49] (MSP, containing physician billing records), BC Vital Statistics [50] (BCVS, capturing deaths and underlying causes), BC Provincial Corrections [51] (recording incarceration entries and releases), Perinatal Care Database[52] (PSBC, covering maternal and infant outcomes), National Ambulatory Care Reporting System[53] (NACRS, documenting Emergency Department visits), Client Roster[54] (providing demographic and geographic data), and the BC Social Development and Poverty Reduction database[55] (tracking social assistance). These datasets are linked at the individual level using a de-identified personal health number assigned by the BC Ministry of Health.[56] A description of these databases and the timeline for data extraction is detailed in **S2 Table**.

### Study population

We will consider ten sub-populations, based on their prior experience with OAT, in the target trial emulations to identify people who achieve better outcomes from SROM compared to methadone and buprenorphine/naloxone. In each case, the study population will include adults (≥18 years) and exclude those who are pregnant or incarcerated at OAT initiation, and individuals with a history of cancer or palliative care. OAT episodes will be defined as continuous treatment with no gaps exceeding four days for methadone and SROM or five days for buprenorphine/naloxone (see **Fig. 1** for details).

**Fig 1.**
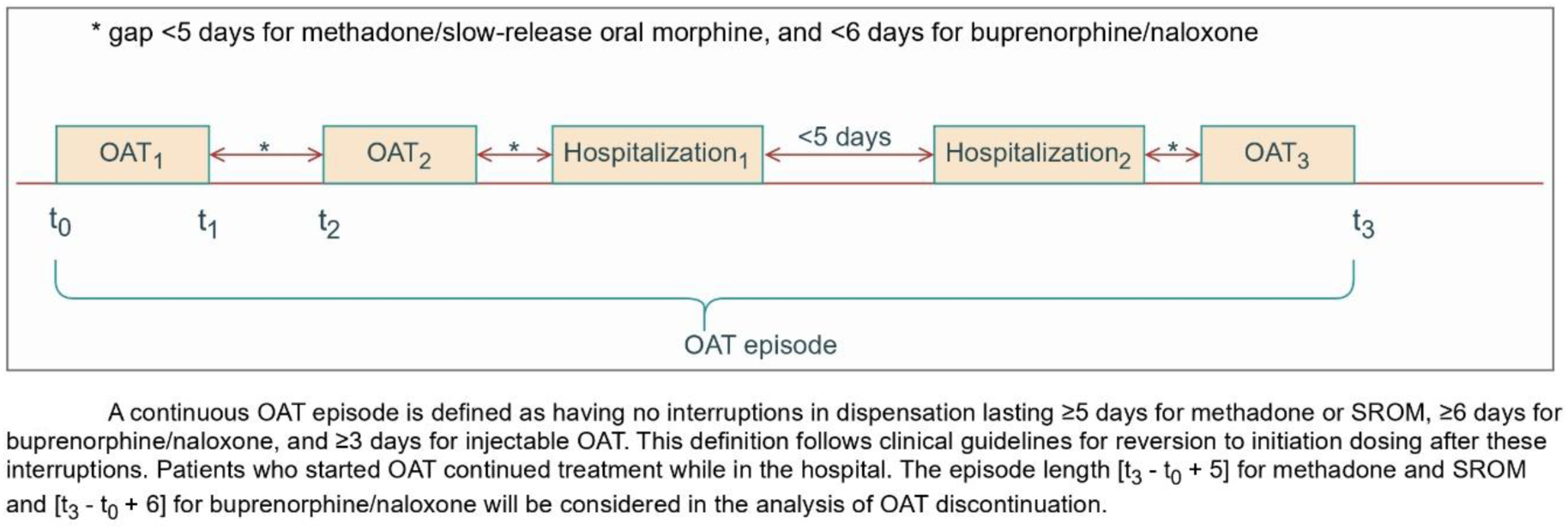
Construction of treatment episode for opioid agonist treatment for comparing slow-release oral morphine versus methadone and buprenorphine/naloxone

These thresholds align with BC guidelines recommending re-titration after these durations.[6] Key sub-populations include (i) incident users – no history of OAT dispensation dating back to January 1, 1996, (ii) prevalent new users – no dispensation for ≥5 days of methadone and SROM and ≥6 days for buprenorphine/naloxone, intended to capture repeated treatment attempts. (iii) prevalent new users – a history of methadone dispensation within past year of the index OAT episode. (iv) prevalent new users – a history of buprenorphine/naloxone dispensation within past year of the index OAT episode, (v) prevalent new users – a history of methadone and buprenorphine/naloxone dispensation within past two years of the index OAT episode, (vi) prevalent new users – completed methadone induction within past two year of the index OAT episode. Induction completion is defined as no dose change within at least two consecutive weeks, (vii) prevalent new users – completed buprenorphine/naloxone induction within past two year of the index OAT episode, (viii) prevalent new users – completed methadone and buprenorphine/naloxone induction during past five years of the index OAT episode, (ix) prevalent new users – individuals initiated treatment with buprenorphine/naloxone, (x) prevalent new users – individuals initiated treatment with methadone (see **Table 2** for details).

**Table 2:** Target study populations for comparative effectiveness analysis.

| Study population | Definition | Rationale |
| --- | --- | --- |
| Incident users | (i) No history of OAT dispensation, dating back to January 1, 1996 | To evaluate treatment outcomes among individuals initiating OAT for the first time |
| Prevalent new users | (ii) No dispensation for $\geq 5$ days of methadone and SROM and $\geq 6$ days for buprenorphine/naloxone | To capture repeated OAT attempts after a washout period, reflecting common real-world patterns. |
| Prevalent new users with past OAT experience | (iii) A history of methadone dispensation within past year of the index OAT episode | To assess individuals with recent prior experience of specific OAT which may influence current treatment response. |
|  | (iv) A history of buprenorphine/naloxone dispensation within past year of the index OAT episode |  |
|  | (v) A history of methadone and buprenorphine/naloxone dispensation in past two years of the index OAT episode |  |
| Prevalent new users who had previously completed OAT induction* | (vi) Completed methadone induction within past two year of the index OAT episode | To assess individuals recently stabilized on methadone and buprenorphine/naloxone, which may influence response to new treatment. |
|  | (vii) Completed buprenorphine/naloxone induction within past two year of the index OAT episode |  |
|  | (viii) Completed methadone and buprenorphine/naloxone induction in past five years of the index OAT episode | A history of completing both inductions may reflect complex needs or higher tolerance, informing comparative effectiveness. |
| Individuals initiated treatment with buprenorphine/naloxone | (ix) Individuals initiated OAT with buprenorphine/naloxone will be included. Switching to SROM within 30 days of initiation will be compared with staying on buprenorphine/naloxone in a two-arm study. | Early switching within buprenorphine/naloxone episode may indicate suboptimal response or poor tolerability. |
| Individuals initiated treatment with methadone | (x) Individuals initiated OAT with methadone will be included. Switching to SROM within 30 days of initiation will be compared with staying on methadone in a two-arm study. | Early switching within methadone episode likely reflects adverse effects such as sedation, emotional blunting, or QT-related cardiac risk. |

\* Induction completion is defined as no dose change within at least two consecutive weeks.

Pregnant and incarcerated individuals will be excluded since pregnancy requires unique clinical management for treatment interruptions,[6] and incarceration involves distinct treatment protocols.[57]

### Key measures

For sub-populations (i) to (viii), the primary exposure is the initiation of OAT with the dispensation of SROM, methadone and buprenorphine/naloxone in an initiator analysis framework. To compare the treatments at key dosing benchmarks, we will follow individuals receiving at least one dispensation of the following dosages per week. The key dosing benchmarks are aligned with BC clinical guidelines and previous literature:[6,58]

> **Week 1:** SROM (≤240 mg), methadone (≤40 mg), and buprenorphine/naloxone (≤16 mg/4 mg).
>
> **Weeks 2–3:** SROM (≥140 mg), methadone (≥30 mg), and buprenorphine/naloxone (≥12 mg/3 mg).
>
> **Week 4 and beyond:** SROM (≥480 mg), methadone (≥80 mg), and buprenorphine/naloxone (≥16 mg/4 mg).

For sub-populations (ix) and (x), two treatment groups in per-protocol analysis framework will be defined as follows: individuals who switch to SROM within 30 days of initiating OAT with buprenorphine/naloxone or methadone, respectively, versus those who remain on their initial OAT type. Due to the lack of inpatient information on OAT dosing in our linked database, we will assume OAT receipt prior to hospitalization is continued daily throughout the duration of hospitalization.[6] Time to OAT discontinuation and all-cause mortality will be primary outcomes of interest for the initiator analysis. OAT discontinuation is defined as no OAT dispensation for ≥5 days for methadone and SROM, and ≥6 days for buprenorphine/naloxone. Additionally, time to overdose-related acute care visits (identified using ICD-10 codes T40, T42.4, and T43.6) will be assessed as a secondary outcome. The per-protocol analysis will assess the same set of outcomes: time to all-cause mortality, overdose-related acute care visits and OAT discontinuation.

### Study follow-up

The time zero is the first dispensation date of methadone, buprenorphine/naloxone, or SROM. For initiator analysis, follow-up ends at the earliest of outcome, death, loss to follow-up, 24 months after time zero, or December 31, 2022. For per-protocol analysis, we will censor at additional events: (i) treatment switch or falls OAT dose below the key benchmark, and (ii) individuals initiate taper or violate eligibility criteria, such as being pregnant, or incarcerated, or had received cancer or palliative care (see **Table 3**).

**Table 3.** Censoring indication in the primary and secondary analyses for comparing slow-release oral morphine versus methadone and buprenorphine/naloxone.

|  | OAT discontinuation |  | All-cause mortality |  | Overdose-related acute-care visits |  |
| --- | --- | --- | --- | --- | --- | --- |
|  | Initiator | Per protocol | Initiator | Per protocol | Initiator | Per protocol |
| OAT discontinuation | outcome | outcome |  |  |  |  |
| End of study <sup>a</sup> | C<sub>1</sub> | C<sub>1</sub> | C<sub>1</sub> | C<sub>1</sub> | C<sub>1</sub> | C<sub>1</sub> |
| Death | C<sub>2</sub> | C<sub>2</sub> | outcome | outcome | C<sub>2</sub> | C<sub>2</sub> |
| Loss to follow-up | C<sub>2</sub> | C<sub>2</sub> | C<sub>2</sub> | C<sub>2</sub> | C<sub>2</sub> | C<sub>2</sub> |
| Deviation from inclusion/exclusion criteria |  | C<sub>2</sub> |  | C<sub>2</sub> |  | C<sub>2</sub> |
| Deviation from the protocol |  | C<sub>3</sub> |  | C<sub>3</sub> |  | C<sub>3</sub> |
| Treatment switch |  | C<sub>3</sub> |  | C<sub>3</sub> |  | C<sub>3</sub> |
| The overdose-related first acute-care visit |  |  |  |  | outcome | outcome |

However, for sub-populations (ix) and (x), where treatment is considered a time-varying exposure and individuals may be consistent with both treatment strategies up to 30 days of OAT episode, we will apply the ‘clone-censor-weight’ approach to emulate the per-protocol target trial.[59,60] In this approach, each individual will be replicated into two clones, one for each treatment strategy. When individuals are no longer consistent with the assigned strategy due to a treatment switch, taper initiation, or violation of eligibility, we will artificially censor the corresponding clone at that time. For the clone assigned to the switch to SROM group, individuals will remain uncensored if they switch to SROM within 30 days of OAT episode, and be censored at day 30 if they have not switched by then (see **Table 3**).

### Covariate selection

We propose to include all selected confounders identified in a recent methadone versus buprenorphine/naloxone study,[61] which applied a modified disjunctive cause criterion[62] to the augmented list of covariates from systematic reviews.[63,64] These covariates include socio-demographic factors, measures of disease severity and comorbidity, and OAT-related factors. Additionally, COVID-19-related covariates will be included to control potential selection bias introduced by pandemic-related service disruptions and differential treatment accessibility (see **Table 4** for the list of covariates).

**Table 4.** Time fixed and time-varying covariates to estimate the inverse probability of treatment and censoring weights.

| Variable | Category | How the variable is defined and measured |
| --- | --- | --- |
| *Socio-demographic factors* |  |  |
| Age | Continuous | Based on birth date information from the client registry. |
| Sex | Male/Female | Based on medical records that record sex. |
| Living in rural region | Yes/No | Based on the population of the assigned local health area[65] |
| Homelessness/unstable housing in past 5 years<sup>‡</sup> | Past year, Past 1-5 years, Never/>5 years | Defined through International Classification of Diseases (ICD) 10: Z59.0, Z59.1 from physician billing or hospital discharge records or 3 consecutive records of no fixed address from Social Development and Poverty Reduction (SDPR) |
| Incarcerated in past year<sup>‡</sup> | Yes/No | Admission to BC correctional facilities and identified through records in the past year from the provincial corrections database. |
| Receipt of income assistance in the past year<sup>‡</sup> | Yes/No | Identified through records in SDPR and PharmaCare plan information on medication dispensations, using records within the past year. |
| Calendar year of time zero | 2010 to 2022 | Determined based on the dispensation date from PharmaNet. |
| *Comorbidity* |  |  |
| Severe mental disorder (ever)<sup>‡, a</sup> | Yes/No | Includes major depressive disorder, bipolar disorder, and schizophrenia, and identified through ICD codes if diagnosed ever and updated if diagnosed during episode follow-up. |
| Alcohol use disorder (ever)<sup>‡, a</sup> | Yes/No | Identified through ICD codes and DIN/PIN, if diagnosed ever and updated if diagnosed during episode follow-up. |
| Hepatitis C virus (ever)<sup>‡, a</sup> | Yes/No | Identified through ICD codes if diagnosed ever and updated if diagnosed during episode follow-up. |
| Asthma (ever)<sup>‡, a</sup> | Yes/No | Identified through ICD codes* and DIN/PIN within the past year, updated during the follow-up. |
| Chronic pain in past year<sup>‡, a</sup> | Yes/No | Identified through ICD codes within the past year, updated during follow-up. |
| Chronic obstructive pulmonary disease<sup>‡, a</sup> | Yes/No | Identified through ICD codes within the past year, updated during the follow-up. |
| Charlson comorbidity index in past year<sup>‡</sup> | 0 and above | Using the Charlson comorbidity index[66] and hospitalization records within the past year, and updated during the follow-up. |
| Dispensations of psychiatric or sedative medication in the past month<sup>‡, a</sup> | Yes/No | Identified through DIN/PIN of dispensations in the past 30. Updated during the follow-up. |

| *Factors related to OAT experience* |  |  |
| --- | --- | --- |
| Dispensations opioid other than OAT in the past month<sup>‡, a</sup> | Yes/No | Identified through DIN/PIN and a case-finding algorithm used prior[67] and updated during the follow-up. |
| Received prescription opioids other than OAT prior to OUD diagnosis<sup>a</sup> | Yes/No | Identified through DIN/PIN and a case-finding algorithm used prior[67] and updated during the follow-up. |
| Treated tobacco use disorder in past year<sup>‡, a</sup> | Yes/No | Based on ICD-10 codes and indication of nicotine replacement therapy in PharmaNet |
| Attachment to the prescriber <sup>‡, a</sup> | Yes/No | Identified based on billing history from the physician. More than 50% of physician billing attributed to the prescriber in the past 12 months |
| Funding of prescribers who initiated OAT: alternative payment plan | Yes/No | Alternative payment plan indicated by the absence of a medical services plan billing record (capturing fee-for-service billing) at initial OAT dispensation. |
| Urine drug test in the past week<sup>‡</sup> | Yes/No | Receipt of urine drug test from physician billing records |
| Practice size<sup>‡</sup> | Large, medium, small | Determined by network analysis; number of unique OAT clients in past 12months. Large for >300 clients, medium for 16-300 clients and small for ≤15 clients. |
| *COVID-19 related factors* |  |  |
| Virtual care<sup>‡</sup> | Yes/No | Virtual care indicated in physician billing records.[68,69] |
| OAT pharmacy delivery<sup>‡</sup> | Yes/No | Indication of delivery of OAT in PharmaNet |
| Receipt of prescriber safer supply (PSS) <sup>‡</sup> | Yes/No | Concurrent PSS dispensations in PharmaNet.[70] |

DIN: drug identification number
PIN: product identification number;
<sup>‡</sup> Time dependent variable will be updated weekly;
a. ICD codes for each condition and DIN/PINs for different medications can be found elsewhere.[58]

All time-varying covariates will be updated weekly. These covariates will be used to estimate the inverse probability of treatment as well as censoring weights to adjust for measured confounding and informative censoring respectively.

### Statistical analysis

We will adjust for all baseline covariates to estimate the initiator effect as hazard ratios, differences of cumulative incidence (risk) at 24 month, and risk curves using a marginal structural Cox proportional hazard regression model (MSM) with inverse probability of treatment weighting.[71,72]. For the per-protocol analysis, additional time varying confounding will be adjusted via censoring weights to mitigate selection bias from artificial censoring due to receipt of dose below key benchmark, treatment switching, violation of eligibility criteria including initiating pregnancy, incarceration, or treatment taper. However in the ‘clone-censor-weight’ approach for the sub-population (ix) and (x), we will not artificially censor for the violation of dosing benchmark and will estimate the artificial censoring weights conditional on their covariate history. Sandwich variance estimates will be used to compute the 95% compatibility (confidence) intervals for the adjusted hazard ratios,[73] while 500 bootstrap samples (with replacement) would be used to compute the compatibility intervals for risk differences and compatibility curves for risk curves.[74] We will apply high-dimensional propensity score (hdPS) and instrumental variable (IV) approaches for the initiator analysis as sensitivity analyses.[75,76] While the MSM and ‘clone-censor-weight’ approaches will target the marginal treatment effect, the IV analysis will estimate a local average treatment effect among compliers, i.e., individuals who would be prescribed the treatment preferred by treating physician or facility (see details in IV estimation section later). We will use SAS version 9.4 to prepare analytic data and conduct the statistical analysis using the SAS and R version 4.5.0.

### Initiator analysis

We will apply a multinomial logistic model to estimate the probability of initiating SROM, methadone or buprenorphine/naloxone, incorporating selected baseline covariates, to adjust for confounding by indication at time zero. The treatment weights will be estimated by taking the inverse of the probability of initiating the observed form of OAT. The selection bias induced by censoring due to death or loss to follow-up will be controlled via censoring weights estimated from a pooled logistic regression model.^67^ To estimate weekly censoring weights, the model will incorporate an indicator for week (modelled with cubic splines), product of week and medication, and both time-fixed and time-varying covariates ().

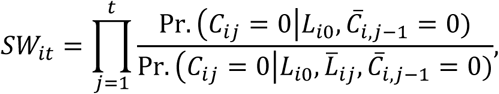

where *SW_it_* is the stabilized weight for the *i*th individual at time *t*, *C_ij_* is the censoring status for the *i*th individual at *j*th time, *C̄_ij_* is the censoring history for the *i*th individual at *j*th time, *L*_*i*0_ is the list of time-fixed covariates including treatment arm, *L̄_ij_* is the time-dependent covariate history for the *i*th individual at *j*th time. Time-dependent covariates will be lagged by one week to ensure censoring is predicted using prior-week covariates. Same set of baseline and time-varying covariates will be included in the treatment and censoring models. Both treatment weights at time zero and censoring weights will be stabilized, truncated at the 99th percentile to limit the influence of extreme values and their product will be used as the final weights.[72] Covariate balance will be assessed using standardized mean differences (SMD), with SMD <0.10 considered balanced.[77] To fit the marginal structural model, a pooled weighted logistic regression model will be fitted to compare the time to OAT discontinuation, all-cause mortality, and overdose-related acute care visits across treatment groups. Based on the fitted models, individual time-to-event (survival) probabilities will be predicted weekly for each treatment arm by setting the treatment equal to a specific value (e.g., SROM, methadone, or buprenorphine/naloxone) for all individuals. These predicted probabilities will then be averaged across the population to get week-specific survival probabilities. The risk of the events will be estimated as one minus the averaged survival probability, with risk differences calculated for SROM initiators versus buprenorphine/naloxone and methadone. We will report hazard ratios, risk differences, and risk curves. Compatibility intervals and curves will be computed using the approach described in the Statistical analysis section.

### hdPS estimation

As a sensitivity analysis, the treatment weights in the initiator analysis will be estimated using hdPS approach, an alternative to propensity score weighting, to address potential residual confounding, particularly for unmeasured differences in the severity of OUD and medical comorbidities among treatment groups. This semi-automated, data-driven method will identify proxy variables from administrative data for inclusion in propensity score models.[75] We will extract covariates from five data sources: DAD diagnostic codes (ICD-10, 4 digits), DAD procedure codes (5 digits), MSP diagnostic codes (ICD-9, 3 digits), MSP fee codes (5 digits), and PharmaNet drug therapeutic class 5 code (8 digits). Proxy variables will be assessed within a six-month window before time zero with exclusions for variables related to investigator-selected covariates, exposure, outcomes, colliders, and known instruments. Proxy covariates appearing in <50 individuals will be excluded to avoid concerns of positivity violations. To refine selection, three recurrence variables will be created for each candidate proxy (any occurrence, frequent occurrence above the median, and frequent occurrence above the 75th percentile), with duplicates removed. A Cox-LASSO regression will rank and select the top 200 covariates per outcome.[78] The selected empirical covariates, combined with investigator-selected covariates, will be incorporated into propensity score models estimated using multinomial logistic regression models or penalized regression methods such as LASSO to estimate the treatment weights.[79] In the marginal structural model, we will apply pooled weighted logistic regression using the final weights—calculated as the product of hdPS weights and censoring weights from the initiator analysis—to estimate hazard ratios, risk differences, and risk curves. Compatibility intervals and curves will be computed using the approach described earlier.

### IV estimation

We will apply IV estimation for initiator analysis as a sensitivity method to address potential uncontrolled confounding. Given the lack of established IV methods for multi-category exposures in time-to-event analyses, we will define a binary exposure: SROM versus either methadone or buprenorphine/naloxone. Two candidate IVs will be tested: (a) prescriber-level preference for SROM and (b) facility-level preference for SROM, both measured among OAT clients in the six months prior to time zero.[80] The validity of the IV relies on four key assumptions. First, the relevance assumption will be tested using a logistic regression model to assess the association between the IVs and the exposure while adjusting for individual- and prescriber-level characteristics. The exclusion restriction and exchangeability cannot be directly verified, so their plausibility will be assessed through expert clinical input from our scientific advisory committee. Additionally, we will evaluate bias due to omitted measured covariates by comparing non-IV and IV analyses.[81] The assumption of homogeneity defines whether the IV-exposure association remains consistent across measured confounders of the exposure-outcome association.[82] The monotonicity assumption requires that there are no “defiers”, i.e., individuals who would always receive opposite treatment to that assigned by IV, and is expected to hold trivially in this context since OAT initiation with SROM is largely determined by prescriber or facility preference rather than individual choice. Since homogeneity and monotonicity cannot be empirically tested, their validity will be assessed through expert consultation.[80,83] To estimate the local average treatment effect, we will apply a two-stage residual inclusion (2SRI) approach.[84,85] In the first stage, a logistic regression approach will model the exposure as a function of the IV and selected covariates, generating residuals that account for uncontrolled confounding. These residuals will be incorporated into a Cox-PH model alongside the exposure and selected covariates. From this fitted model, we will estimate hazard ratios, risk differences, and risk curves. Compatibility intervals and curves will be computed using the approach described earlier.

### Per-protocol analysis

We will conduct a per-protocol analysis to estimate treatment effects under sustained adherence at key dosing benchmarks, complementing the initiator effect.[86] To control the selection bias due to artificial censoring, we will consider two censoring models along with the propensity score models: (i) accounting for treatment switching, receipt of dose below key benchmark, and (ii) accounting for death, loss to follow-up, protocol violation by initiating pregnancy, dose taper, incarceration, or receipt of cancer or palliative care (see **Table 3** for outcome specific censoring mechanism). Dose tapering is defined as two consecutive daily dose reductions without an intervening increase. Both censoring weights will be estimated using the approach described in the Initiator analysis section earlier. Final weights will be the product of the stabilized inverse probability of treatment weights at time zero and the post-baseline inverse probability of censoring weights, truncated at the 99th percentile. In the marginal structural model, we will apply a pooled logistic regression approach incorporating the final weights. The fitted model will be used to compute hazard ratios, risk differences, and risk curves. Compatibility intervals and curves will be computed using the approach described earlier. For sub-populations (ix) and (x), the censor-weighting approach will be similar, but we will apply a modified censoring mechanism as described in **Table 3**.

### Subgroup and sensitivity analyses

We will conduct a range of sensitivity analyses by restricting the study population and timeline to pre-COVID era, redefining key variables, and applying alternative estimation methods including hdPS estimation, IV estimation (see **Table 5** for details).

**Table 5:**
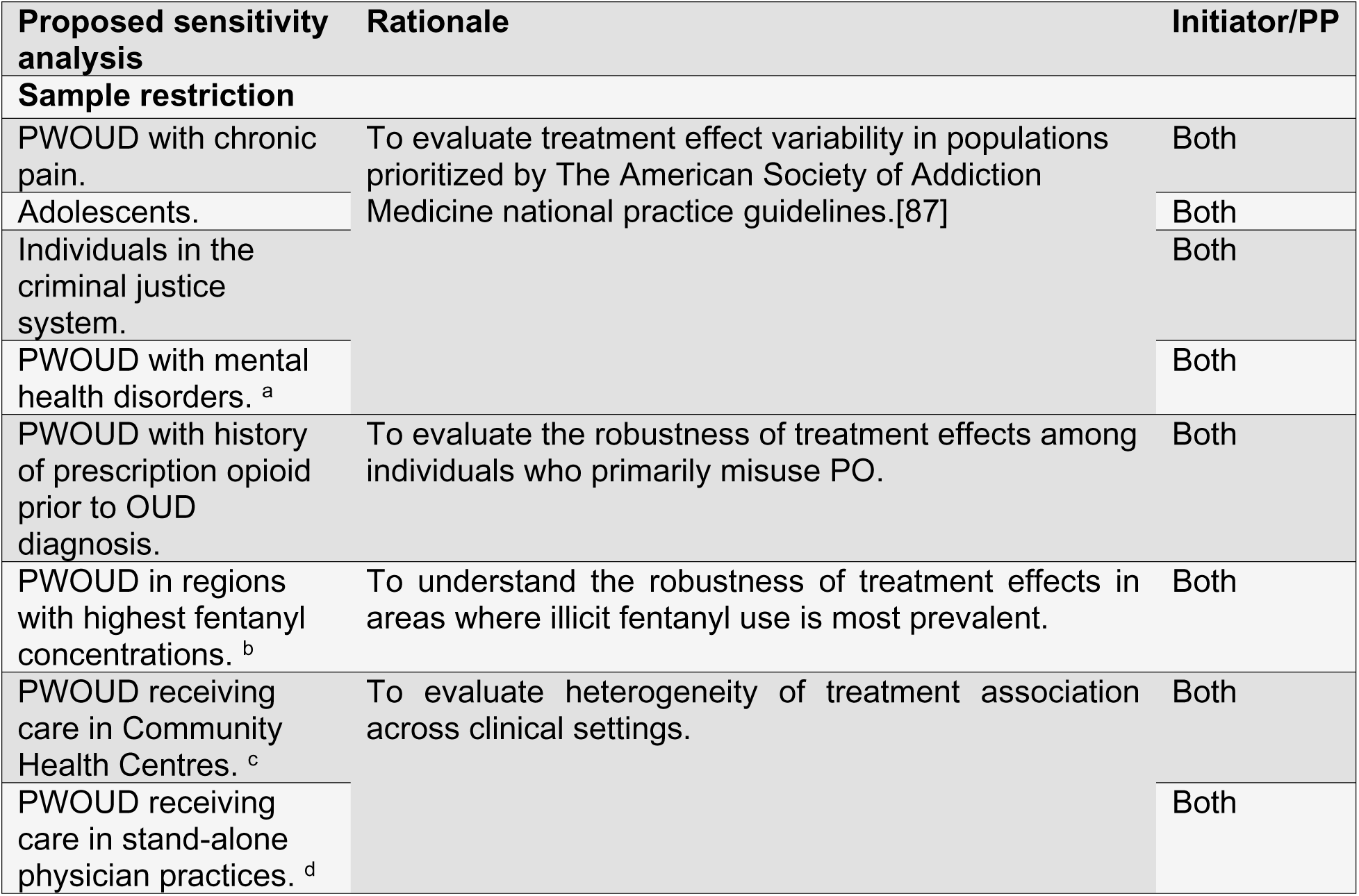
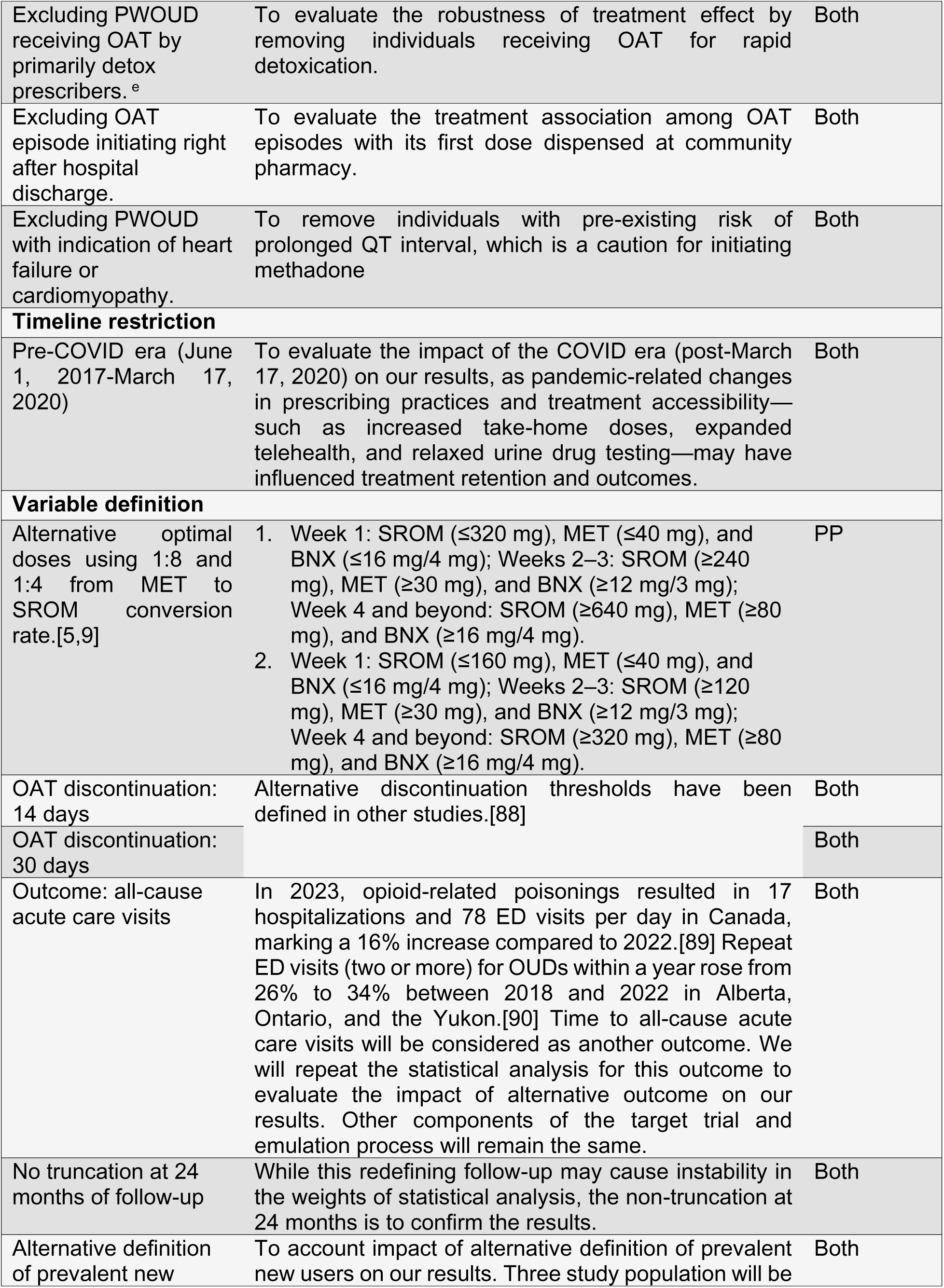
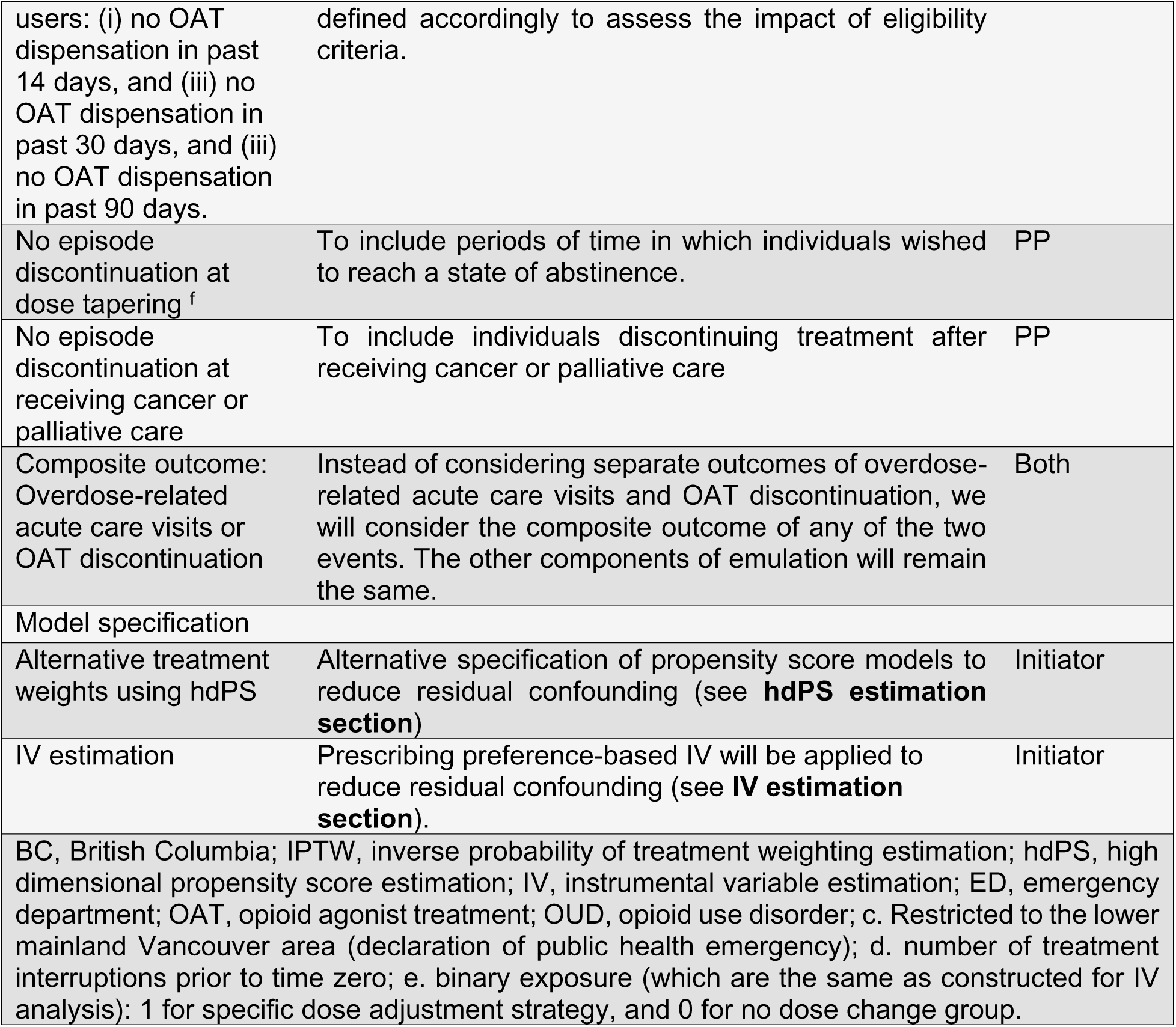
Proposed subgroup and sensitivity analysis.

Because fentanyl had already become the dominant opioid in the drug supply by 2017, it will not be feasible to examine the effect of pre-fentanyl era on our findings. Relevant findings from each analytical approach will be presented in a forest plot. Any deviations from this protocol will be documented in the final reports.

### Ethics and Dissemination

This study uses de-identified administrative health data accessed under data-sharing agreements with the BC Ministry of Health and the Ministry of Mental Health and Addictions. Data were fully de-identified before being provided to the research team; no identifiable personal information was accessed. This initiative is categorized as a quality improvement project. The Providence Health Care Research Institute and the Simon Fraser University Office of Research Ethics reviewed the project, deeming it exempt according to Article 2.5 of the 2018 Tri-Council Policy Statement: Ethical Conduct for Research Involving Humans.[91] Because the study was conducted using de-identified administrative data and involved no direct contact with individuals, the requirement for informed consent was waived by the reviewing bodies. No participants, including minors, were contacted, and no written or verbal consent was sought or required. This study adheres to international guidelines, including Strengthening the Reporting of Observational Studies in Epidemiology guidelines,[92] and will be administered for ex post evaluation by a multidisciplinary scientific advisory committee. The results will be disseminated through peer-reviewed journals both electronically and in print, as well as at national and international conferences. This research aims to generate robust evidence for the effectiveness of SROM versus methadone and buprenorphine/naloxone dependent on one’s prior OAT experience, with the goal of improving[93] and life-saving[94] medications.

### Study status and timeline

The de-identified administrative health data from June 1, 2017 to December 31, 2022 has been obtained, but no analyses have been conducted to date. Data cleaning, cohort construction, and all planned analyses will begin following protocol approval and the study is expected to be completed within six months afterwards.

## Discussion

This study will generate population-level evidence on the comparative effectiveness of SROM, methadone, and buprenorphine/naloxone across ten clinically relevant sub-populations of both new and experienced OAT users. By emulating target trials with linked administrative data, we will estimate both the initiator and per-protocol effects of these medications. The application of marginal structural models, along with multiple sensitivity analyses, is designed to mitigate confounding by indication and other systematic biases that commonly affect observational OAT research. Findings will inform evidence-based refinement of OAT guidelines and support clinical decision-making in BC and other jurisdictions experiencing rising opioid-related harms.

A key limitation is the inability to directly measure several determinants of OAT selection, such as OUD severity, in-hospital OAT dispensing, non-medically attended overdoses, medication allergies or hypersensitivities, and changes in mental health symptoms, because they are not fully captured in administrative datasets. To reduce the resulting risk of residual confounding, we will use proxy measures available in administrative data in hdPS adjustments. Despite these constraints, the large population coverage, rigorous design, and comprehensive sensitivity analyses will provide robust and policy-relevant evidence on the comparative effectiveness of SROM relative to methadone and buprenorphine/naloxone.

### Patient and public involvement

Although no patients were directly engaged in designing this study, its conceptualization was shaped by previous interactions with local advocacy organizations representing individuals who use drugs and those accessing OAT.[95] Qualitative feedback on this and other related objectives outlined in the parent grant R01DA050629 was incorporated to prioritize this analysis, taking into account its potential impact on client engagement. The results will be disseminated to local advocacy groups after the analysis is completed.

## Data Availability

No datasets were generated or analysed during the current study. All relevant data from this study will be made available upon study completion.

## Author Contributions

BN conceptualized and secured funding for the study. MHM conducted literature reviews, wrote the first draft of the article and key methodological components of the article. JEM and BN aided in methodological development. All authors provided critical revisions to the manuscript and approved the final draft. MHM is responsible for the overall content as the guarantor.

## Funding statement

This study was funded by the National Institutes on Drug Abuse (NIDA grant no. R01DA050629).

## Notes

### Competing Interest Statement

The authors have declared no competing interest.

### Funding Statement

Yes

### Author Declarations

This study uses de-identified administrative health data accessed under data-sharing agreements with the BC Ministry of Health and the Ministry of Mental Health and Addictions. Data were fully de-identified before being provided to the research team no identifiable personal information was accessed. This initiative is categorized as a quality improvement project. The Providence Health Care Research Institute and the Simon Fraser University Office of Research Ethics reviewed the project, deeming it exempt according to Article 2.5 of the 2018 Tri-Council Policy Statement: Ethical Conduct for Research Involving Humans. Because the study was conducted using de-identified administrative data and involved no direct contact with individuals, the requirement for informed consent was waived by the reviewing bodies. No participants, including minors, were contacted, and no written or verbal consent was sought or required. For further details, please contact

